# Time-variant strategies for optimizing the performance of non-pharmaceutical interventions (NPIs) in protecting lives and livelihoods during the COVID-19 pandemic

**DOI:** 10.1101/2020.04.13.20063248

**Authors:** Yap Wei Aun, Dhesi Baha Raja

## Abstract

A stochastic Individual Contact Model (ICM) using SIR compartments allowing for time-variant parameters was used to simulate 100 non-pharmaceutical intervention (NPI) strategies and exit trajectories for a hypothetical population, and to collect epidemiological and non-epidemiological outcomes to measure the performance of these strategies over the course of a period of intervention (up to six months) for a total duration of one-year to allow the full implications of the strategy and endgame to manifest. We find that variations in the time dimension and intensity of various strategies can have vastly different performance outcomes: (i) the timing of NPIs can ‘shrink the area under the curve’ (cumulative infections) not just ‘flatten the curve’; (ii) prolonged lockdowns have diminishing margins of returns; (iii) smooth, submaximal lockdowns perform better than pulsatile lockdowns; and (iv) the efficiency of various strategies incorporating both epidemiological and non-epidemiological outcomes vary substantially. Most sobering, none of the simulated strategies allow for an ‘acceptable’ path to exit within six months due to very large gaps in health system capacity.

## Introduction

Many countries throughout the world have instituted strong non-pharmaceutical interventions (NPIs), such as lockdowns, in immediate response to rapid increases in COVID-19 cases to slow the contagion so that the health system can cope. As COVID-19 cases moderate where lockdowns are in place, there is debate about how these lockdowns can be withdrawn but inadequate evidence on the performance of various NPI exit strategies to inform these. Epidemiological considerations are critical but given the impact of NPIs on livelihoods and economies, both epidemiological outcomes and non-epidemiological outcomes will need to be considered in the performance of NPIs. This is the critical gap in evidence that this paper aspires to contribute towards by using a modeling platform to simulate 100 different scenarios on a hypothetical population whereby time-variant parameters related to lockdown interventions are modified individually to draw generalizable insights and highlight further knowledge and analytic gaps. Performance measures are collected during the simulation to generate evidence for and against different strategies. NPIs are inextricably linked to exit strategies and hence all simulations are run for an additional 6 months to achieve steady state so that the whole strategy until the endgame can be measured.

At time of writing there is no proven vaccine or other medical breakthrough in the prevention or treatment of COVID-19. Societies are hence faced with a difficult choice between two broad paths. The first path is to strongly suppress the outbreak and eliminate local transmission, leaving the population relatively unravaged by COVID-19 and its associated morbidity and mortality, but fully susceptible to SARS-CoV-2. NPI measures required to attain and preserve this paradigm are substantive and are not short term. They will have major implications on society, the economy, and other aspects of health such as mental health and emergency management of noncommunicable diseases. If a proven and accessible vaccine then becomes available, population herd immunity can be developed through immunization rather than infection. This first path is not the focus of this paper as an exit from NPI is antithetical. The second path leads to a population which is no longer susceptible to SARS-CoV2 through the attainment of herd immunity by infection, implying that a large proportion of the population will be exposed to COVID-19 morbidity and mortality on-route to this destination. The goal of NPIs in this context is to keep the burden of COVID-19 cases within the capacity of the health system. However, not all NPIs are created equal in terms of performance against epidemiological outcomes measures and non-epidemiological outcomes, as explored through simulations in this paper.

## Methods

### Model

We assessed the efficacy of ‘lockdown’-style NPIs using a stochastic Individual Contact Model (ICM) using modified SIR compartments – Susceptible (S) individuals; potentially infectious asymptomatic and symptomatic Infected (I) individuals; and Recovered (R) individuals – for a hypothetical initial closed population comprising 100,000 individuals with population and modeling parameters as described in Table 1. As a closed population, background arrivals (births or immigration) and departures (non-COVID-19 deaths) are not simulated. The SIR model was modified to incorporate the infectivity of exposed individuals who are asymptomatic.

**Table 1:** Initial population and modeling parameters

| Parameter | Value |
| --- | --- |
| Initial population size | 100,000 (Closed population) |
| Initial asymptomatic and symptomatic infected individuals – compartment “I” | 1% |
| Proportion asymptomatic | 0.75 (upper bound) (Carl Heneghan, Jon Brassey, and Tom Jefferson 2020) |
| Mean duration of latent asymptomatic and asymptomatic period | 18 days (Eichenbaum, Rebelo, and Trabandt 2020) |
| Baseline ‘acts’ per person per day | 5 |
| - Reduction in ‘acts’ per person per day with a maximal (100%) lockdown | 68.5%<sup>4</sup> (Google n.d.) |
| Infection probability per ‘act’ | 0.03 |
| Implied R0 | 2.7 |
| Deaths among symptomatic | 1.5% |
| Proportion needing hospitalization | 0.19 (Wu and McGoogan 2020) |
| Duration of hospitalization among those needing hospitalization | 13 (Guan et al. 2020) |
| Number of simulations per scenario | 5 |

### Simulations

For each scenario being modelled, stochastic simulations were performed 5 times and the mean value for each variable of interest for each time period (one day) was taken as the result and 95% confidence intervals generated. Simulations were performed using *EpiModel* version 1.7.5 (Jenness, SM, and Morris 2018) on *R* version 3.6.3 (R Core Team 2020). Modifications to the *EpiModel* package were required to enable time variant parameters for ICM simulations. The source code used for the simulations including modifications made to the *EpiModel* package are published^3^ under the GNU General Public License v3.0.

### Time-variant intervention

The key NPI in these models is a reduction in ‘acts’ per day due to a ‘lockdown’. These reductions can be varied on a day-to-day basis throughout the simulation. Acts represent opportunities to spread infection. Each opportunity is associated with a fixed infection probability per ‘act’.

### Scenarios Simulated

A total of 100 scenarios in 5 series were simulated. Baseline series A comprises a single scenario (A) representing the baseline progression of COVID-19 in this hypothetical population without any NPIs. B-series scenarios simulate a once-off lockdown for a fixed period of 42 days (6 weeks) with variation only to the starting day of the lockdown from Day 2 to Day 72. C-series scenarios simulate a once-off lockdown starting on a fixed day - Day 14 – but varying by duration for 50, 60, 70, 80, 90, 100, 110, and 120 days. D-series scenarios simulate pulsatile ‘on and off’ lockdown strategies as per Table 2 below. This series of scenarios assume that when the lockdowns are in the ‘off’ period of the pulse, there is no rebound increase in acts. The differences in socioeconomic activities and in infection dynamics between weekdays and weekends are not modelled.

**Table 2:** D-series ‘pulsatile’ scenarios

| D-series scenario | Form of pulsatile lockdown | Implied duty-cycle (percent) |
| --- | --- | --- |
| D1a | 2-day work week (no lockdown) followed by a lockdown for the remainder of the week | 71% |
| D1b | 3-day work week (no lockdown) followed by a lockdown for the remainder of the week | 57% |
| D1c | 4-day work week (no lockdown) followed by a lockdown for the remainder of the week | 43% |
| D1d | 5-day work week (no lockdown) followed by a lockdown for the remainder of the week | 29% |
| D2 | One-week of lockdown followed by one-week of no lockdown | 50% |
| D3 | Two-week of lockdown followed by two-week of no lockdown | 50% |
| D4 | Three-week of lockdown followed by three-week of no lockdown | 50% |
| D5 | Four-week of lockdown followed by four-week of no lockdown | 50% |
| D6 | Five-week of lockdown followed by five-week of no lockdown | 50% |
| D7 | Six-week of lockdown followed by six-week of no lockdown | 50% |

E-series scenarios simulate a prolonged period of NPIs all starting on Day 14 and ending on Day 182, but with varying intensity in the reduction of acts per day, with 100 percent representing the maximal lockdown intervention used in scenarios series B to D and 0 percent representing no lockdown.

For all series and scenarios, the simulations were continued for a total duration of 365 days to allow for a resurgence in cases and for a steady state to be reached.

### Outcomes Measures

#### Epidemiological outcomes

Epidemiological outcome measures and non-epidemiological proxy measures are simulated. Epidemiological outcome measures are: (i) Percent of the initial population ultimately infected on Day 365, (ii) deaths per 1,000, (iii) peak prevalence and incidence rates, (iv) days to peak prevalence and incidence. Increased deaths due to an overwhelmed health system are not counted under the deaths per 1,000 outcome measure but are modelled using a proxy measure which is the sum of the squares of daily prevalence throughout the simulation period that is above a health system capacity threshold. The health system capacity threshold is based on the world average hospital beds per capita (World Development Indicators 2020) and adjusted for the mean duration of hospitalization, the proportion of symptomatic “I” cases, the proportion of symptomatic patients requiring hospitalization. The daily prevalence above this threshold was squared to penalize extreme breaches of this day-to-day capacity threshold. This measure of an overwhelmed health system is referred to here as the ‘over-capacity factor’.

#### Non-epidemiological outcomes

Non-epidemiological proxy measures of the of NPIs on economic and social activity are: (i) aggregate acts per person over the intervention period and (ii) aggregate daily acts per person, square rooted, over the intervention period. The square rooted measure prizes smooth rather than pulsatile lockdowns and may be relevant for certain economic and social activities.

### Robustness checks

As this is a stochastic simulation, larger initial population sizes may alter outcome measures. As a robustness check, we ran a selection of simulations with population sizes of 1 million and 10 million. Duration to peak and to attain a steady state are increased by small amounts for larger initial population sizes, but the insights from the simulations remain the same.

Due to limitations of computing capacity available, each scenario is simulated 5 times with the same parameters but differing random draws. Given the initial population of 100,000 and 5 simulations, the standard deviation of epidemiological parameters at each time period over the 5 simulations are modest and are included in the plots. As a robustness check, we ran a selection of scenarios with 10 and 20 simulations. Results remain essentially unchanged except at the margin or at tipping point scenarios.

### Limitations

A key limitation which is relevant to the results, many of which demonstrate ‘tipping point’ phenomena, is the wide variation in parameter estimates available in the public domain at this early period during the COVID-19 outbreak. For example, estimates for the proportion of those testing positive who do not have symptoms vary from 5 to 80 percent (Carl Heneghan, Jon Brassey, and Tom Jefferson 2020). Furthermore, parameter estimates are derived from a wide range of countries, with different population, environmental, and climactic conditions. The simulations here can hence only be interpreted as the relative performance of NPI strategies for a hypothetical population for which parameter estimations used are true, although these findings provide qualitative insights which may be generalizable to other populations with similar characteristics and epidemiological parameters.

The population is closed and hence there is no chance of infected arrivals into the population. This is an important consideration once interventions are lifted on Day 182 (or earlier for some scenarios) as the population may not yet have achieved herd immunity. As a robustness check, a seeding event was planned for all simulations where there were an insignificant number of infected individuals remaining after interventions ceased and where steady state without intervention had not been achieved by Day 365. The seeding event would introduce 1000 new infected individuals into the population on Day 366 to observe if an outbreak can be sustained over a further simulated 365 days.

## Results

Outcome measures for series A, B (a selection of the 71 scenarios), C, D, and E scenarios are tabulated in Table 3. B-series scenarios are presented primarily as charts (Figure 3). Prevalence charts for all scenarios are available on online.^5^

**Table 3:** Epidemiological and non-epidemiological outcomes

| Scenario | Percent Infected | Peak Prevalence Day | Peak Prevalence per 1,000 | Peak Incidence Day | Peak Incidence per 1,000 | Deaths per 1,000 | Over-capacity factor | Acts per person | Acts per person (sqrt) |
| --- | --- | --- | --- | --- | --- | --- | --- | --- | --- |
| Note: ✓ = Best performance within its series |  |  |  |  |  |  |  |  |  |
| A. Baseline (No intervention) | 89.0% | 60 | 230.5 | 47 | 16.9 | 3.1 | 7,757 | 840.00 | 375.66 |
| **B-series: Timing of lockdown** |  |  |  |  |  |  |  |  |  |
| B27. 42-day lockdown starting on Day 27 | 85.0% | 118 | 144.4 | 106 | 10.1 | 3.0 | 5,695 | 701.15 | 336.69 |
| B33. 42-day lockdown starting on Day 33 | 83.1% | 122 | **113.7 ✓** | 34 | 12.4 | 3.0 | 5,220 | 701.15 | 336.69 |
| B34. 42-day lockdown starting on Day 34 | 82.5% | 35 | 118.3 | 35 | 13.5 | 2.8 | 5,127 | 701.15 | 336.69 |
| B47. 42-day lockdown starting on Day 47 | 77.2% | 48 | 195.9 | 46 | 17.2 | 2.8 | 5,436 | 701.15 | 336.69 |
| **C-Series: Duration of lockdown** |  |  |  |  |  |  |  |  |  |
| C1. Duration of lockdown: 50 days | 87.2% | 124 | 189.6 | 112 | 13.7 | **3.0 ✓** | 6,658 | **673.75 ✓** | **328.84 ✓** |
| C2. Duration of lockdown: 60 days | 87.1% | 135 | 185.0 | 120 | 13.5 | 3.1 | 6,541 | 639.50 | 319.03 |
| C3. Duration of lockdown: 70 days | 86.8% | 148 | 178.8 | 133 | 12.8 | **3.0 ✓** | 6,397 | 605.25 | 309.22 |
| C4. Duration of lockdown: 80 days | 86.6% | 162 | 174.7 | 148 | 12.5 | 3.2 | 6,283 | 571.00 | 299.41 |
| C5. Duration of lockdown: 90 days | 86.7% | 173 | 174.5 | 162 | 12.3 | **3.0 ✓** | 6,251 | 536.75 | 289.60 |
| C6. Duration of lockdown: 100 days | 86.3% | 187 | 169.9 | 178 | 12.1 | 3.2 | 6,137 | 502.50 | 279.79 |
| C7. Duration of lockdown: 110 days | 86.3% | 201 | 167.7 | 188 | 12.0 | 3.1 | 6,082 | 468.25 | 269.98 |
| C8. Duration of lockdown: 120 days | **86.2% ✓** | 214 | **165.1 ✓** | 201 | **11.7 ✓** | 3.2 | **6,014 ✓** | 434.00 | 260.17 |
| **D-Series: Pulsatile** |  |  |  |  |  |  |  |  |  |
| D1a. 2-day workweek; 5-day lockdown | 77.2% | 244 | **60.5 ✓** | 78 | **5.5 ✓** | 2.8 | **3,435 ✓** | 434.00 | 260.17 |
| D1b. 3-day workweek; 4-day lockdown | **71.3% ✓** | 85 | 84.6 | 71 | 8.4 | **2.5 ✓** | 3,811 | 516.20 | 283.71 |
| D1c. 4-day workweek; 3-day lockdown | 74.7% | 78 | 123.8 | 64 | 11.3 | 2.8 | 4,982 | 598.40 | 307.26 |
| D1d. 5-day work week 2-day lockdown | 80.6% | 71 | 162.9 | 57 | 13.3 | 2.8 | 6,091 | **680.60 ✓** | **330.80 ✓** |
| D1e. Lockdown alternate weeks | 71.9% | 85 | 108.7 | 71 | 10.2 | 2.7 | 4,400 | 557.30 | 295.48 |
| D2. Lockdown alternate 2-weekly periods | 73.1% | 99 | 115.3 | 71 | 11.2 | **2.5 ✓** | 4,357 | 557.30 | 295.48 |
| D3. Lockdown alternate 3-weekly periods | 74.0% | 99 | 122.2 | 57 | 10.2 | 2.7 | 4,285 | 557.30 | 295.48 |
| D4. Lockdown alternate 4-weekly periods | 74.7% | 71 | 116.9 | 71 | 12.4 | 2.6 | 4,323 | 557.30 | 295.48 |
| D5. Lockdown alternate 5-weekly periods | 72.3% | 85 | 143.5 | 85 | 13.6 | 2.7 | 4,391 | 509.35 | 281.75 |
| D6. Lockdown alternate 6-weekly periods | 76.3% | 99 | 162.8 | 99 | 14.1 | 2.8 | 4,691 | 557.30 | 295.48 |
| **E-series: Intensity variation** |  |  |  |  |  |  |  |  |  |
| E1.90% lockdown | 83.5% | 266 | 130.6 | 254 | 8.8 | 3.0 | 5,040 | 327.14 | 234.87 |
| E2. 80% lockdown | 80.1% | 255 | 88.2 | 236 | 5.8 | 2.8 | 3,938 | 384.68 | 254.80 |
| E3. 70% lockdown | 75.4% | 83 | **52.0 ✓** | 15 | **3.9 ✓** | **2.5 ✓** | **3,315 ✓** | 442.22 | 273.26 |
| E4. 60% lockdown | **71.1% ✓** | 89 | 75.6 | 72 | 4.7 | **2.5 ✓** | 3,660 | 499.76 | 290.54 |
| E6. 50% lockdown | 72.0% | 81 | 102.2 | 74 | 6.6 | **2.5 ✓** | 4,485 | 557.30 | 306.84 |
| E7. 40% lockdown | 75.8% | 80 | 129.7 | 62 | 8.6 | 2.8 | 5,302 | 614.84 | 322.32 |
| E8. 30% lockdown | 80.3% | 72 | 158.3 | 57 | 10.9 | 2.9 | 6,063 | 672.38 | 337.08 |
| E9. 20% lockdown | 83.7% | 67 | 182.2 | 53 | 13.0 | 3.0 | 6,683 | 729.92 | 351.22 |
| E10. 10% lockdown | 86.6% | 64 | 208.0 | 50 | 15.1 | 2.9 | 7,272 | **787.46 ✓** | **364.80 ✓** |

**Figure 1:**
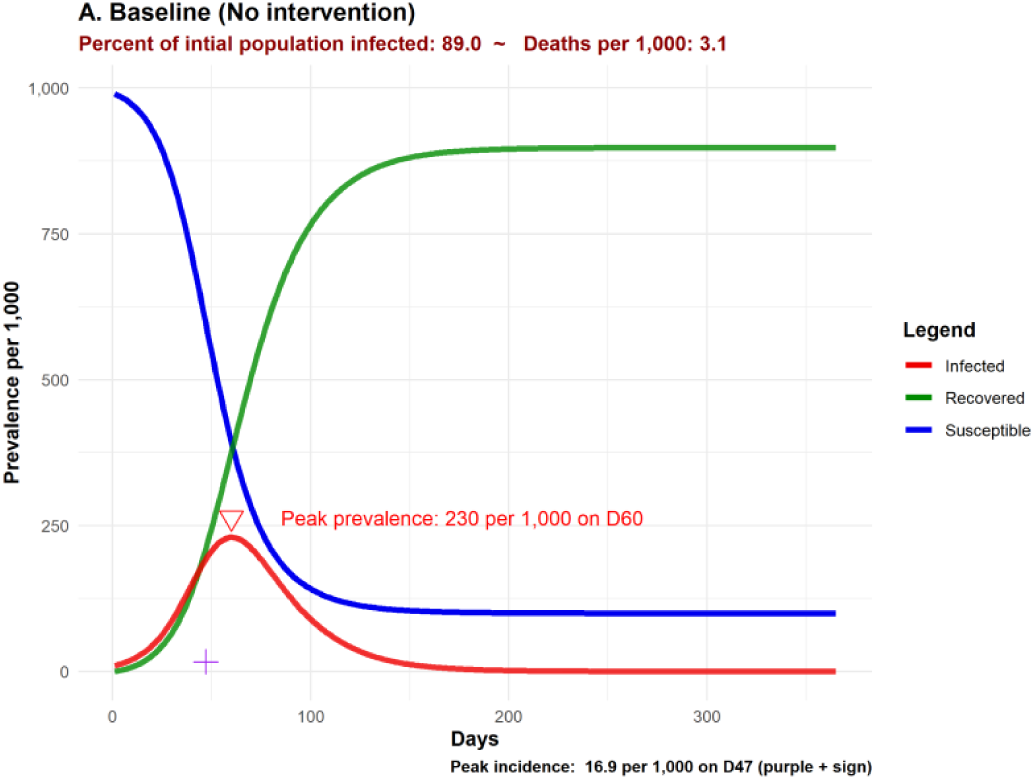
Baseline Scenario A prevalence plot

**Figure 2:**
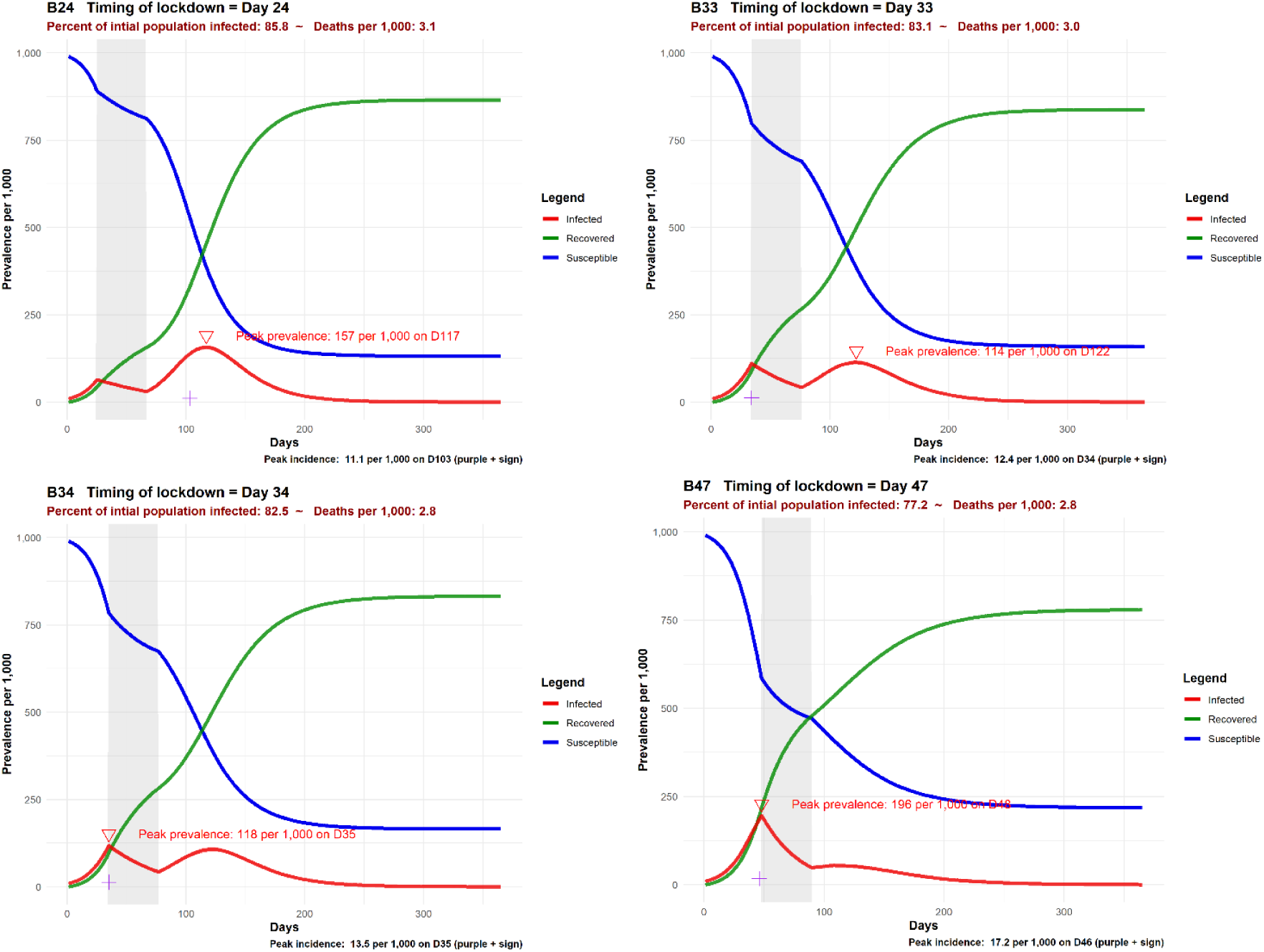
Selected B-series prevalence plots

**Figure 3a and b:**
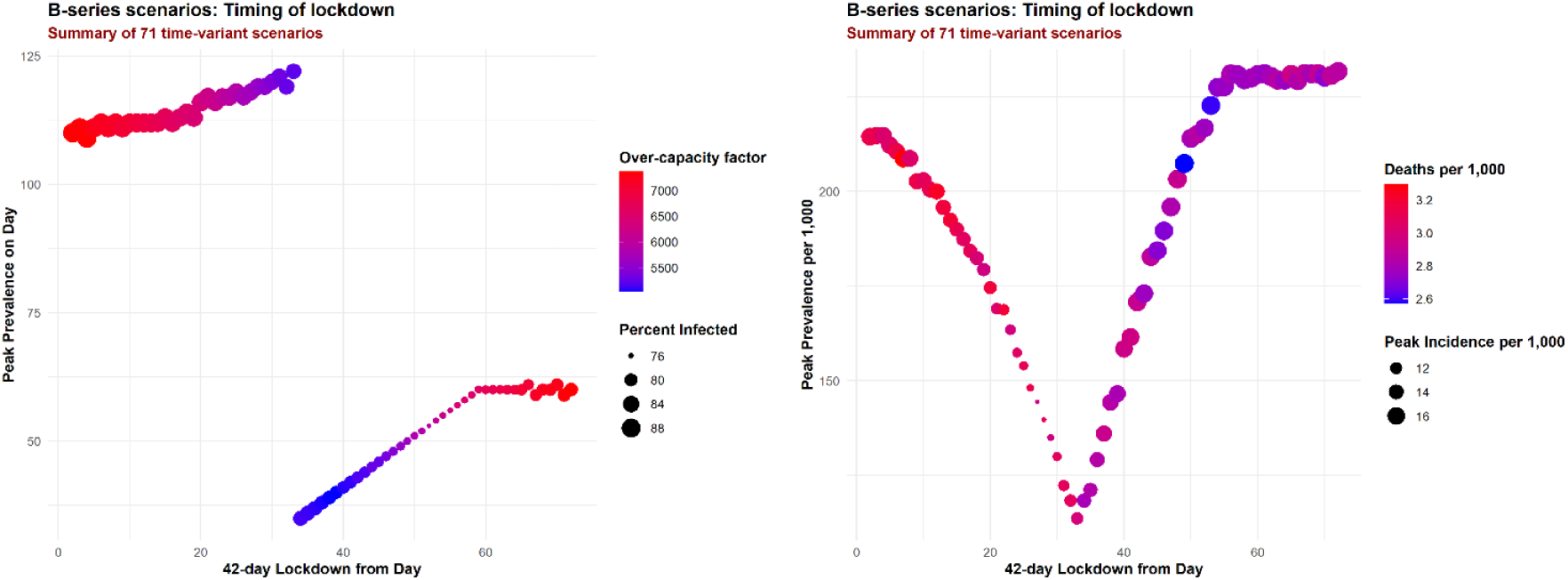
B-series scenario performance

### Baseline scenario

The baseline scenario devoid of any NPI simulated has a peak prevalence of 231 per 1,000 on Day 60 and a peak incidence of 16.9 per 1,000 on Day 47. A total of 3.1 deaths per 1,000 are simulated by Day 365, excluding excess deaths resulting from an overwhelmed health system, as 88.5 percent of the initial population is infected in this fast spreading outbreak. The over-capacity factor is 7,757, the highest of all scenarios, reflecting an unmitigated surge in COVID-19 cases overwhelming the health system. However, proxy measures of economic and social activity are not reduced by any NPIs and hence, in reflection of business-as-usual, are the highest of all scenarios at 840 cumulative ‘acts’ per person over the intervention period from Day 14 to 182.

### Timing of introduction

The performance of B-series simulations varies by the day the fixed-duration lockdown is introduced (Figure 2 and Figure 3) and demonstrates clearly how bimodal peaks in prevalence can be created by a once-off lockdown. Very early introduction of the lockdown merely delays the outbreak until after the lockdown but a lockdown introduced a bit later (B24) reduces peak prevalence to 157 per 1,000 compared to the baseline as a substantial number of cases occur around the introduction of and during the lockdown in a relatively moderated manner. Late introduction of the lockdown (B47) performs poorly in outcomes such as reducing or delaying peak prevalence but performs well in reducing the final tally of the infected to just 77.2 percent of the initial population. The best performance in terms of reducing peak prevalence is seen when the lockdown is timed to result in roughly equally high bimodal peaks, at a critical tipping point such that just one day’s difference in timing the lockdown results in the highest peak flipping from occurring after the lockdown on Day 122 (B33) to occurring one day after the lockdown is introduced Day 35 (B34). This tipping point is seen most vividly in Figure 3a where the day of peak prevalence shifts dramatically. Although this critical point results in the lowest peak prevalence Figure 3b, the lowest over-capacity factors are seen in scenarios where the lockdown was introduced a few days after this critical point. Scenarios which result in the lowest peak incidence are those timed before this critical point and scenarios timed many days after this critical point have the best performance in reducing deaths (excluding excess deaths due to an overwhelmed health system).

### Duration of lockdown

C-series simulations results indicate than for all the durations of lockdown simulated, ranging from 50 to 120 days, infections were *not* eradicated from the population and the population remained susceptible to COVID-19 at the end of the lockdown. Hence, between 60 and 80 days (the longer lockdowns, the longer the delay) *after* the end of the lockdown, the prevalence of COVID-19 cases peaked following a resurgence, albeit with lower peaks for prevalence and incidence compared with the baseline scenario. The longer the duration of the lockdown, the lower the peaks for prevalence and incidence. However, proxy measures of economic and social activity are severely reduced by prolonged lockdowns. For example, the cumulative ‘acts’ per person for the 120-day lockdown is 434, which is the same as that for a 2-day work week with 5-days of lockdown throughout the intervention period.

### Pulsatile lockdowns

D-series simulation results generally show stronger performance for high-frequency (time varying strategies within a period of a week) but no one D-series scenario demonstrates superiority across all outcome measures. D1a, a 2-day work week followed by 5-days of lockdown has the best performance in reducing the over-capacity factor and peak prevalence and incidence, but not in the final tally of deaths or percent infected due to a bimodal peak which occurs long after the intervention ends on Day 244. D1a also has an implied duty cycle of 71%, the tightest lockdown of all the D-series scenarios. Scenario D1d is notable only as a comparison to the baseline – if a full 5-day work week is allowed, with positive implications on business and industry, but weekends are under lockdown, there are substantial improvements to epidemiological outcomes.

### Smooth, submaximal lockdowns

E-series simulation results demonstrate critical tipping point dynamics and include the best performing scenarios (see Figure 7 Scenario E3 or E4, depending on epidemiological outcome of interest) among all scenarios simulated. For NPIs which are implemented strongly, under the parameter estimates used for this hypothetical population, these would represent lockdowns with an intensity of 80% to 100% (see Figure 7 Scenario E2), the NPI suppresses the outbreak during the intervention period but to an extent that prevents the population from attaining herd immunity. Hence, when lifted, the outbreak peaks rapidly. Weak lockdowns with an intensity of 50% or below do not suppress the outbreak and permit a peak even during the period of lockdown. An unstable equilibrium (E3), exists where the right amount of suppression can flatten out the prevalence curve very well over several months. At the end of the intervention, the right balance of immunity within the community and remaining infections fester at a low intensity for a few months more before dissipating without any further NPIs. Importantly, this ‘sweet spot’ is *only relevant* for the parameter estimates used, the population characteristics, the stage of the outbreak at which the lockdown was started and the duration of this simulated partial lockdown (168 days from Day 14). The scenario conditions for E3 are *not* a general rule-of-thumb which can be applied to other populations.

**Figure 4a and b:**
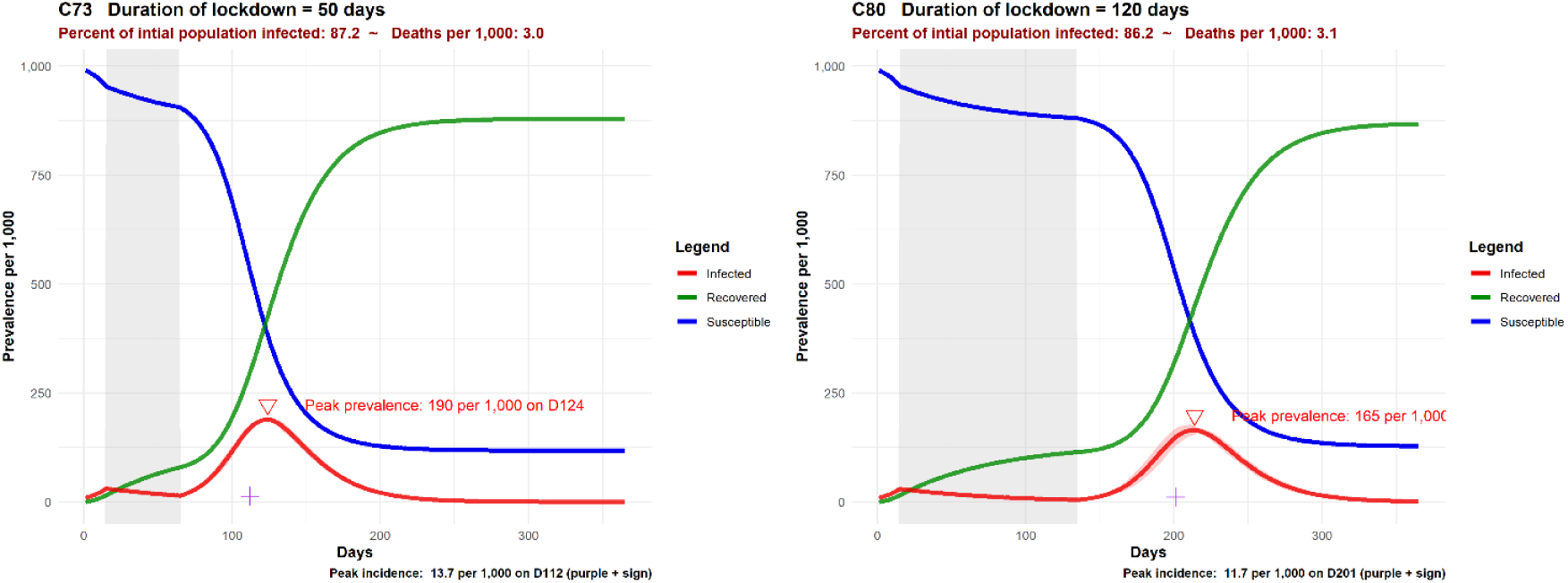
Selected C-series prevalence plots – shortest (4a) and longest (4b) duration

**Figure 5:**
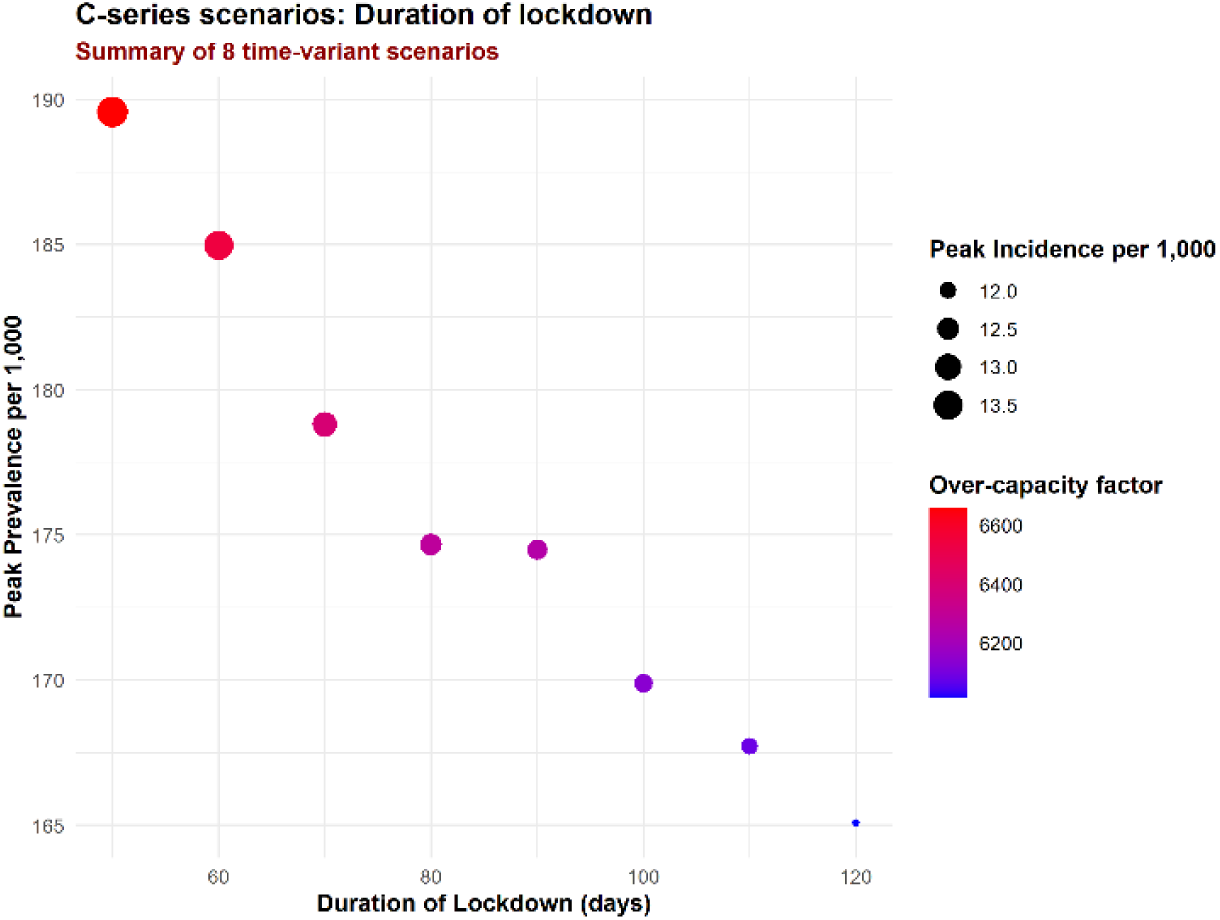
C-series scenario performance

**Figure 6:**
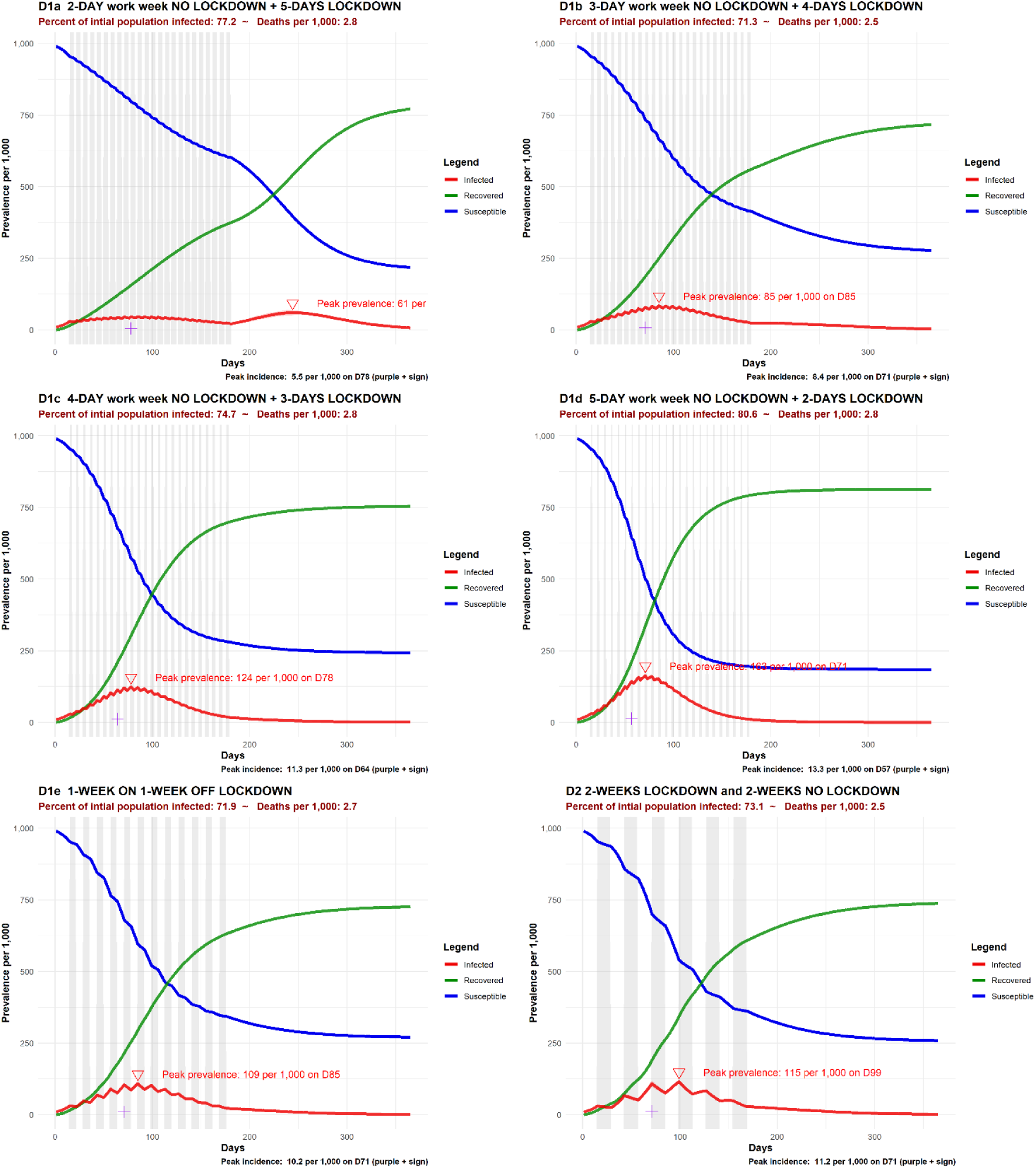
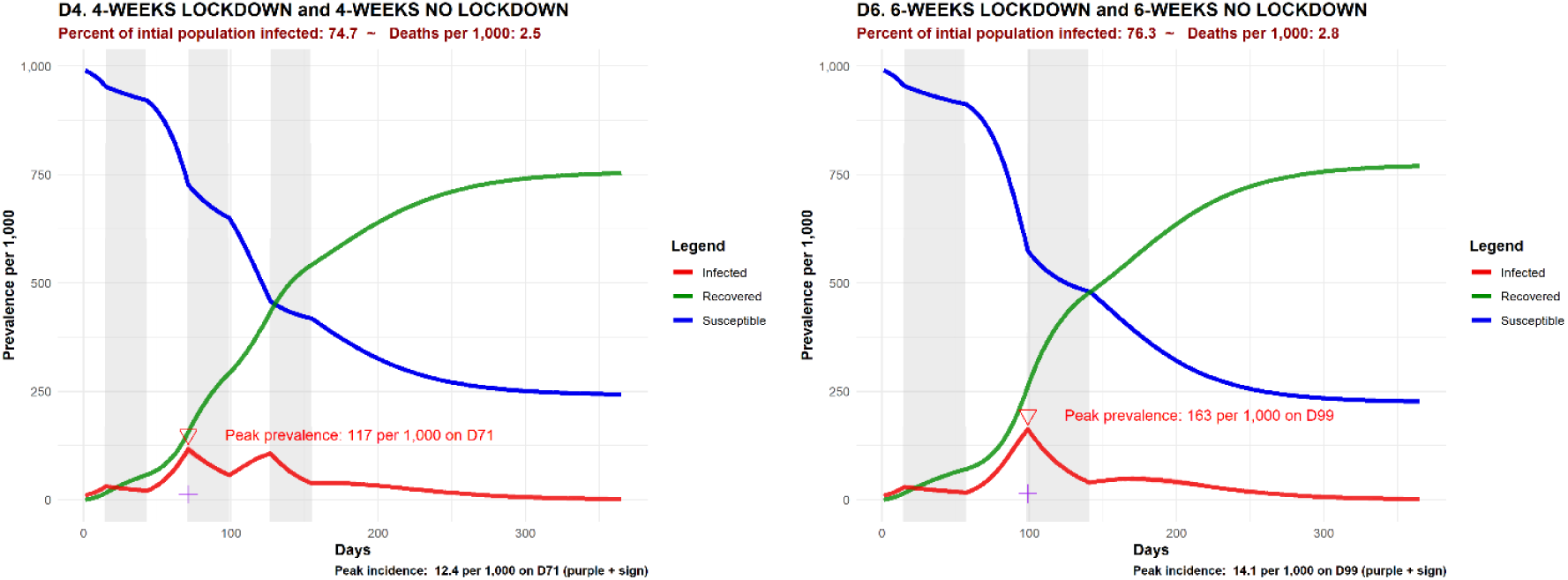
Selected D-series prevalence plots

**Figure 7:**
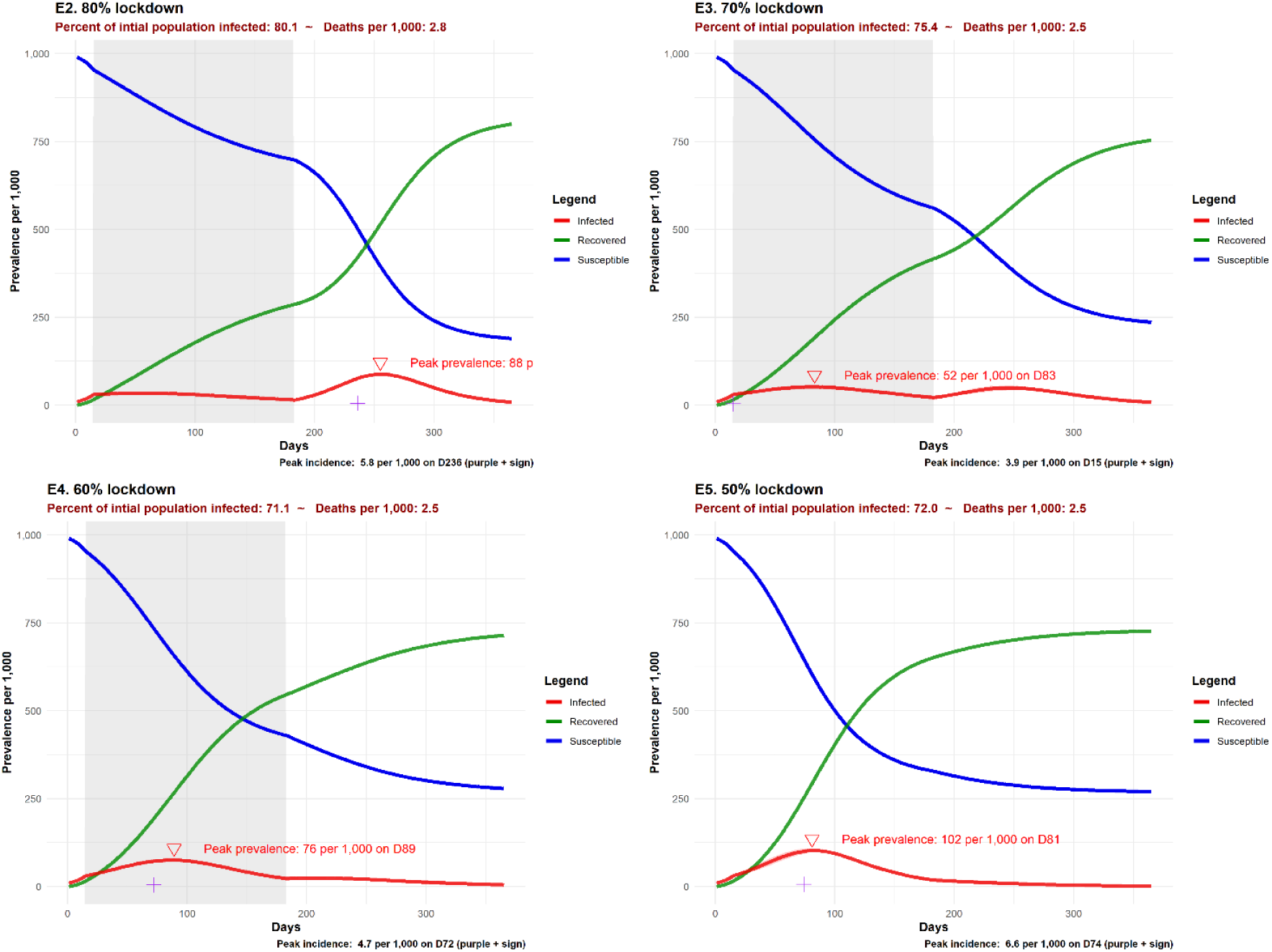
Selected E-series prevalence plots

Scenario E5 is an important comparator against D-series scenarios as the reduction in ‘acts’ is the same as pulsatile scenarios with a 50% duty cycle (i.e., D1e to D6). Peak prevalence and incidence for Scenario E5 are lower than for any of these D-series comparators.

### Steady state

In all scenarios, a steady state was achieved by Day 365, despite the presence of remaining infected individuals at the end of interventions on Day 182. Note that for some scenarios, such as a premature lifting of a lockdown followed by no further measures, a progression similar to the baseline scenarios whereby a large outbreak peak occurred followed by a natural dissipation due to herd immunity was attained. None of the scenarios hence trigger the requirement for seeding with infected individuals as described in methodology.

## Discussion

### ‘Shrink the area under curve’ not just ‘flatten the curve’

In all scenarios simulated^6^, the percentage of the initial population infected by Day 365, at which steady state without prior elimination of infections and without NPIs has been reached (analogous to herd immunity), is high but varies widely by scenario. In the baseline scenario, this percentage is the highest at 89.0 percent. In the most optimal scenario simulated for reducing cumulative infections (Figure 7 Scenario E4) this was 17.9 percentage points lower. Applied on a global scale, this corresponds to sparing 1.4 billion people from the morbidity and mortality associated with COVID-19 infection. B-series scenarios indicate that NPIs not only flatten the curve to avoid overwhelming health system capacity but shrink the area under the curve especially when applied towards the end of the outbreak. This insight is relevant even for countries which may have missed instituting NPI measures early as a final punch of NPIs to prevent overshoot, even after cases has peaked, will likely reduce mortality and morbidity *independently* of the effect of keeping the burden of COVID-19 cases within health system capacity.

### Timing affects different dimensions performance

If a single lockdown is envisaged, this should not be done too early or too late. For best reductions in peak prevalence, the timing should aim to ‘split the mountain’ into two roughly equal bimodal peaks. For best reductions in cumulative infections, covering the later stage of the outbreak with a later lockdown is superior, but at the expense of higher peak prevalence. This point may already be moot for many countries which have instituted lockdowns in response to a spike in COVID-19 cases but may be issues to be considered for those not yet under lockdown.

### Diminishing marginal returns for extending prolonged lockdowns

The extra 70 days of a 120-day of lockdown, compared with a 50-day lockdown, merely delayed the peak day of incidence by 20 days and did not prevent a resurgence of COVID-19 as the population remains non-immune after the intervention. Under the epidemiological parameters and population characteristics simulated for this hypothetical population, none of the C-series ‘smash-the-mountain’ strategies were able to eradicate COVID-19 fully, even in this closed population of 100,000. The technical strength of the stochastic ICM model used for these simulations, compared to deterministic compartment models, is that full eradication is a possible outcome as individuals are modelled discretely. Unless a vaccine is expected, a prolonged lockdown with an intensity much greater than that required to keep COVID-19 cases within health system capacity, but which is not able to eliminate the infection, is of unclear value. These C-series scenarios are among the worst performing strategies.

### Smooth, submaximal lockdowns are more effective than pulsatile lockdowns, in terms of epidemiological outcomes and non-epidemiological outcomes

In terms of epidemiological outcomes, the best performing scenario (E3 or E4) is a smooth and constant partial lockdown applied throughout the intervention period. For any given duty cycle (the fraction of one period that a pulsatile lockdown is in force), the equivalent smooth partial lockdown was superior (also see Kissler et al. 2020). A potential caveat to this would be if certain social or economic activities price the chance, even if short, to be fully ‘active’. Under the non-epidemiological outcomes simulated in this, we include a linear measure (‘acts’ per person) which is equivocal to the difference between working at half intensity all the time vs working at full intensity half the time, and a non-linear measure (square root of ‘acts’ per person) which discounts the need to work at full intensity for at least some short periods. Low-frequency lockdowns with periods longer than two-weeks had notably poor performance, most likely due to long ‘off’ periods whereby NPI measures are removed.

Submaximal lockdowns have an unstable equilibrium and can tip in either direction resulting in diminished performance. This inherent instability makes it challenging for any party to be able to hit the right spot perfectly. However, smooth submaximal and high-frequency lockdown strategies allow for easier real-time calibration of intensity during the intervention. Further analysis would be required to identify the appropriate triggers from empirical surveillance findings on the ground to inform calibration during the intervention. Submaximal lockdowns do not need to have a fixed intensity throughout and the authors conjecture that a positively skewed gentle hill with a final bump to terminate local transmissions as the population approaches the minimum level of herd immunity may be ideal. Identifying this curved bimodal lockdown and calibration strategies is an area for further research.

### Pushing the frontiers of efficiency

Modeling both epidemiological and non-epidemiological outcomes allows a notion of efficiency to be constructed, even without making any trade-offs between these two outcomes. Figure 9 plots an epidemiological outcome (peak prevalence) on the y-axis and a non-epidemiological outcome (the square root of the sum of daily ‘acts’ per person) on the x-axis. For scenarios within the shaded area, another alternative scenario can be found which is superior from either an epidemiological and/or non-epidemiological perspective. Inferior scenarios can hence be discarded from consideration unless there are other outcome dimensions not captured in these two variables which are relevant. The most optimal scenarios are at the boundary of the shaded area – the ‘efficiency frontier’. Further analysis should be done to generate scenarios superior to the current frontier – either iso-epidemiological strategies (with identical epidemiological outcomes) but superior non-epidemiological outcomes (a move to the RIGHT in the figure) and/or strategies which improve epidemiological outcomes without compromising non-epidemiological outcomes (a move to the BOTTOM of the figure).

**Figure 8:**
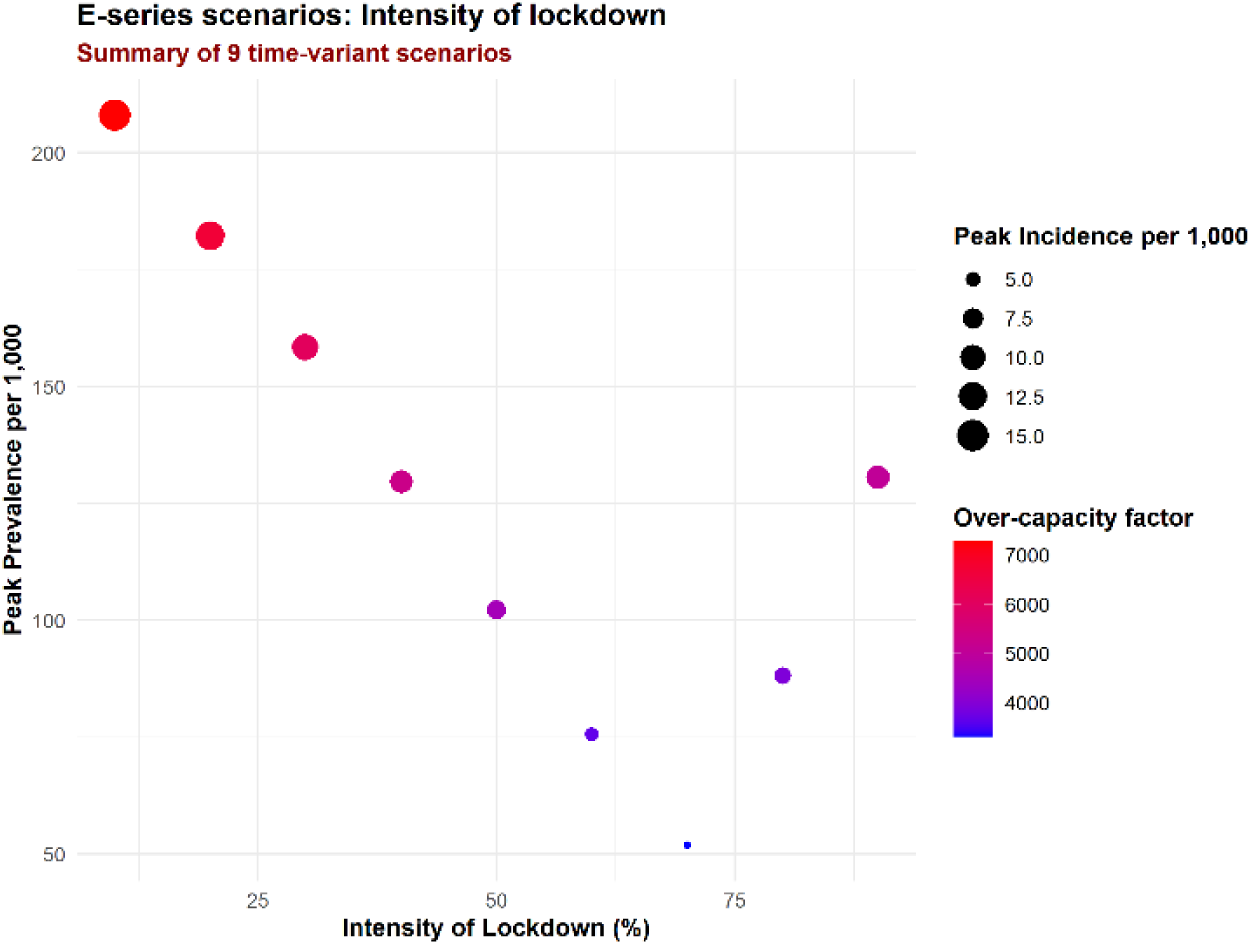
E-series scenario performance

**Figure 9:**
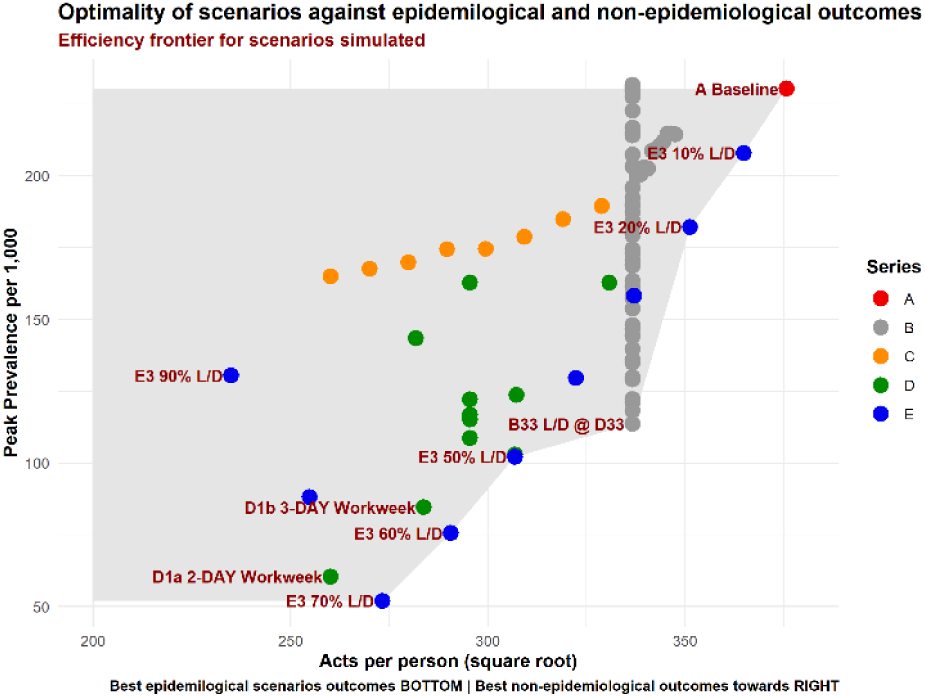
NPIs and efficiency

### No ‘acceptable’ path to exit within 6 months

If acceptability is defined in terms of avoiding overwhelming health system capacity, none of the scenarios modeled allow for an ‘acceptable’ exit within 6 months. An exit is defined in this context as a population which has enough immune individuals to prevent a further outbreak from being sustained without any NPIs in place. The highest performing model flattens the curve substantially from a peak prevalence of 231 per 1,000 in the baseline scenario to 52 per 1,000 (Figure 7 Scenario E3) with six months of intervention **but will still overwhelm the health system capacity by an order of magnitude or more for the 6 months of the intervention followed by 6 months for the outbreak to dissipate on its own**. Further analysis is warranted to devise an optimal path towards an exit which does not overwhelm health system capacity, but the modeling assumptions in scenario E3 can inform the lower bound of the socioeconomic price – an NPI which keeps COVID-19 cases within health system capacity will need to stretch on for longer than 6 months and at greater intensity than the 70% used in this scenario.

## Data Availability

Source codes will be released to https://github.com/quanticlear/Optimizing-NPI-performance

## Acknowledgements

We would like to express gratitude to all first line workers throughout the world who are providing critical services during the COVID-19 pandemic.

## Footnotes

3 https://github.com/quanticlear/Optimizing-NPI-performance

4 Used the mean of four categories: Retail, Grocery, Transit and Work

5 https://github.com/quanticlear/Optimizing-NPI-performance

6 It is highly probable, that a scenario involving a prolonged full lockdown to suppress COVID-19 for most of the duration of the simulation would prevent such infections, but this ‘path one’ strategy (suppress and eliminate local transmission), although perfectly valid, is not the focus of this paper.

